# T cell and autoantibody profiling for primary immune regulatory disorders

**DOI:** 10.1101/2024.02.25.24303331

**Authors:** Emily M. Harris, Sarah Chamseddine, Anne Chu, Leetah Senkpeil, Matthew Nikiciuk, Aleksandra Bourdine, Logan Magin, Amer Al-Musa, Brian Woods, Elif Ozdogan, Sarife Saker, David P. Hoytema van Konijnenburg, Christina S.K. Yee, Ryan W. Nelson, Pui Lee, Olha Halyabar, Rebecca C. Hale, Megan Day-Lewis, Lauren A. Henderson, Alan A. Nguyen, Megan Elkins, Toshiro K. Ohsumi, Maria Gutierrez-Arcelus, Janique M. Peyper, Craig D. Platt, Rachael F. Grace, Brenna LaBere, Janet Chou

## Abstract

**Background:** Limited clinical tools exist for characterizing primary immune regulatory disorders (PIRD), which are often diagnoses of exclusion. Increased CD4^+^CXCR5^+^PD1^+^ circulating T follicular helper (cTfh) cell percentages have been identified as a marker of active disease in some, but not all, autoimmune disorders.

**Objective:** To develop a diagnostic approach that combines measurements of cellular and serologic autoimmunity.

**Methods:** We recruited 71 controls and 101 pediatric patients with PIRD with autoimmunity. Flow cytometry was used to measure CD4^+^CXCR5^+^ T cells expressing the chemokine receptors CXCR3 and/or CCR6. IgG and IgA autoantibodies were quantified in 56 patients and 20 controls using a microarray featuring 1616 full-length, conformationally intact protein antigens. The 97.5^th^ percentile in the controls serves as the upper limit of normal for percentages of cTfh cells, CD4^+^CXCR5^+^ T cells expressing CXCR3 and/or CCR6, and autoantibody intensity and number.

**Results:** We found that 27.7% of patients had increased percentages of CD4^+^CXCR5^+^PD1^+^ cTfh cells and 42.5% had increased percentages of CD4^+^CXCR5^+^ cells expressing CXCR3 and/or CCR6. Patients had significantly more diverse IgG and IgA autoantibodies than controls and 37.5% had increased numbers of high-titer autoantibodies. Integrating measurements of cTfh cells, CD4^+^CXCR5^+^ T cells with CXCR3 and/or CCR6, and numbers of high-titer autoantibodies had 71.4% sensitivity (95% CI: 0.5852 – 0.8158) and 85% specificity (95% CI: 0.6396 – 0.9476) for patients with PIRD compared to controls.

**Conclusion:** By integrating CD4^+^ T cell phenotyping and total burden of autoantibodies, this approach provides additional tools for the diagnosis of PIRD lacking clinical diagnostic criteria.

**Highlights Box:**

- Primary immune regulatory disorders (PIRD) are heterogenous and often diagnoses of exclusion if no genetic cause is identified. Current diagnostic tools do not combine cellular and serologic measures of autoimmunity.
- Measuring activated CD4^+^ T cells expressing the chemokine receptors CXCR3 and/or CCR6 and the total number of circulating autoantibodies can enhance detection of autoimmunity in PIRD beyond the capabilities of currently used tools.
- This study identifies new indicators of autoimmunity that can be feasibly implemented and leveraged for improving the diagnosis of PIRD.

## Introduction

Primary immune regulatory disorders (PIRD) can target any organ, leading to clinical heterogeneity that hinders diagnosis.^1^ Autoimmunity is a key feature of many types of PIRD^1^ Many autoimmune features, such as immune thrombocytopenia (ITP), are diagnoses of exclusion due to limited diagnostic testing, and thus necessitate a “watch and wait” management approach that can delay effective treatment.^2^ Positivity for disease-associated autoantibodies does not reliably correlate with disease activity due to the variable recognition of epitopes inherent in a polyclonal response. Many disease-associated autoantibodies recognize intracellular antigens whose contributions to disease pathogenesis remain unknown; furthermore, most disorders have no associated autoantibodies.^3–5^ Although next-generation DNA sequencing has identified some monogenic causes of autoimmunity, most patients lack a genetic diagnosis.^6^ Delayed diagnoses contribute to disease progression and morbidity through postponed or inadequate treatment. The collective impact of autoimmune disorders worldwide has highlighted the need for expanding diagnostic options for these patients. ^7–10^

Mechanistically, autoimmunity arises from self-reactive T and B cells that generate inflammatory cytokines and autoantibodies. T follicular helper (Tfh) cells are CD4^+^ T cells with robust capacity for inducing B cell activation and class-switching.^11^ Upon recognizing antigen, T cells upregulate the chemokine receptor CXCR5.^12^ While effector T cells downregulate CXCR5 upon terminal differentiation, Tfh cell progenitors maintain expression of this chemokine receptor as it enables their migration to the B cell follicle.^11^ Subsequently, sustained interactions between Tfh and B cells upregulate additional proteins characteristic of Tfh cells in germinal centers: the transcription factor B cell lymphoma 6 protein (Bcl6) and the immune checkpoint receptor, programmed cell death 1 (PD-1).^11,13^ Notably, the periphery contains CD4^+^CXCR5^+^PD1^+^ T cells that lack expression of Bcl6.^14,15^ These cells are commonly referred to as circulating Tfh (cTfh) cells due to features shared with Tfh cells in the germinal center, including T cell receptor clonotypes and the ability to induce class-switching.^14–17^ Based on these T-B cell interactions underlying the generation of autoreactive cells, we previously showed that percentages of CD4^+^CXCR5^+^PD1^+^ cTfh cells exceeding 12% of CD4^+^ T cells have high sensitivity and specificity for identifying rare and common multi-systemic autoimmune disorders, such as activated protein kinase delta syndrome and SLE.^18^ However, increased percentages of cTfh cells are not a universal feature of all PIRD indicating the need for additional diagnostic approaches.^19^ Measurement of cTfh cells also does not account for effector T cells that can home to tissues, secrete proinflammatory cytokines, and damage target organs.

Despite the essential contributions of T and B cells to the development of autoimmune disorders, existing diagnostic tests do not combine measures of cellular and serologic autoimmunity. To address this gap, we used an approach that incorporates: (1) measurement of cTfh cells, (2) the expression of chemokine receptors required for the homing of cTfh and effector T cells to secondary lymphoid organs and end-organ targets, and (3) quantification of total autoantibody burden using a high-throughput protein microarray as an indicator of serologic autoimmunity. We focused on patients with PIRD because these diseases have far fewer diagnostic tools and clinical criteria than common autoimmune diseases. We recruited patients with diverse types of autoimmunity reflecting the reality of clinical practice and focused on measures of cellular and serologic autoimmunity that could be implemented in a clinical care setting.

## Methods

### 2.1 Study participants

Informed consent was obtained from patients, all of whom were recruited at Boston Children’s Hospital. This study was approved by the Institutional Review Board (IRB-04-09-113R) of Boston Children’s Hospital. The study included a cohort of 101 patients and 74 controls (**Table 1**).

**Table 1.**
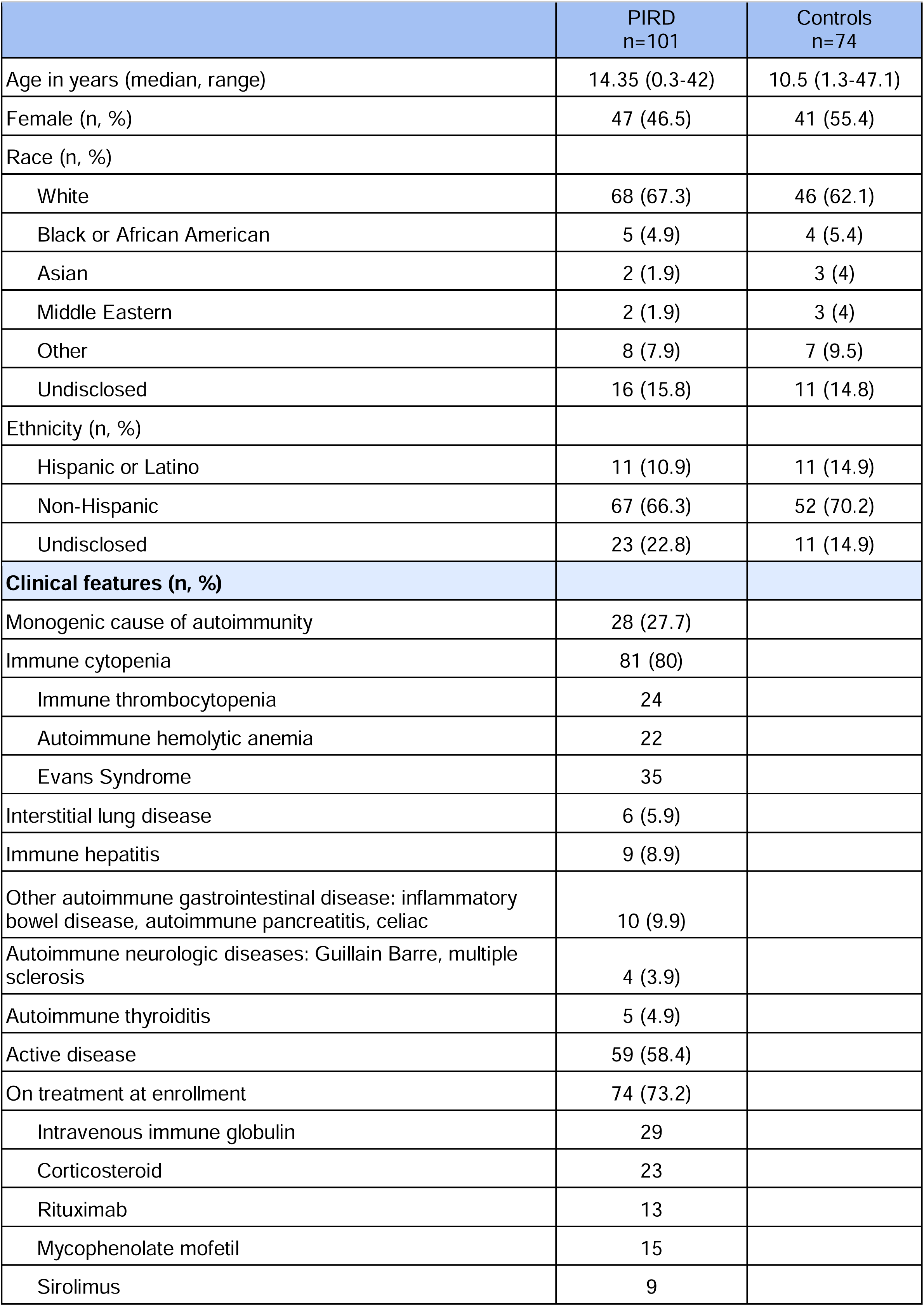
Characteristics of study cohort and controls.

|  | PIRD<br>n=101 | Controls<br>n=74 |
| --- | --- | --- |
| Age in years (median, range) | 14.35 (0.3-42) | 10.5 (1.3-47.1) |
| Female (n, %) | 47 (46.5) | 41 (55.4) |
| Race (n, %) |  |  |
| White | 68 (67.3) | 46 (62.1) |
| Black or African American | 5 (4.9) | 4 (5.4) |
| Asian | 2 (1.9) | 3 (4) |
| Middle Eastern | 2 (1.9) | 3 (4) |
| Other | 8 (7.9) | 7 (9.5) |
| Undisclosed | 16 (15.8) | 11 (14.8) |
| Ethnicity (n, %) |  |  |
| Hispanic or Latino | 11 (10.9) | 11 (14.9) |
| Non-Hispanic | 67 (66.3) | 52 (70.2) |
| Undisclosed | 23 (22.8) | 11 (14.9) |
| **Clinical features (n, %)** |  |  |
| Monogenic cause of autoimmunity | 28 (27.7) |  |
| Immune cytopenia | 81 (80) |  |
| Immune thrombocytopenia | 24 |  |
| Autoimmune hemolytic anemia | 22 |  |
| Evans Syndrome | 35 |  |
| Interstitial lung disease | 6 (5.9) |  |
| Immune hepatitis | 9 (8.9) |  |
| Other autoimmune gastrointestinal disease: inflammatory bowel disease, autoimmune pancreatitis, celiac | 10 (9.9) |  |
| Autoimmune neurologic diseases: Guillain Barre, multiple sclerosis | 4 (3.9) |  |
| Autoimmune thyroiditis | 5 (4.9) |  |
| Active disease | 59 (58.4) |  |
| On treatment at enrollment | 74 (73.2) |  |
| Intravenous immune globulin | 29 |  |
| Corticosteroid | 23 |  |
| Rituximab | 13 |  |
| Mycophenolate mofetil | 15 |  |
| Sirolimus | 9 |  |

Information regarding medical diagnoses and treatments received was obtained through review of the electronic medical record. Race and ethnicity were obtained from self-reported data from the medical record. Controls were recruited from allergy and hematology outpatient clinics where they were evaluated for common conditions such as resolving iron deficiency and allergies not requiring treatment. The control cohort included patients without any history of autoimmunity, immune dysfunction, malignancy, or transplantation. Patients were classified as having a diagnosis of PIRD as defined by the Primary Immune Deficiency Treatment Consortium^20^ and having autoimmunity based on clinical, laboratory, or genetic causes abstracted from the medical record by the study’s clinical hematologists and immunologists prior to measurement of cTfh cells. Patients and controls with any clinical signs or symptoms of acute infection were excluded from the study. Disease activity was defined by abnormal laboratory values or disease-specific symptoms consistent with the respective autoimmune diagnosis. For patients with autoimmune cytopenias, these included a platelet count of less than 150K cells/μL, hemoglobin of less than lower limit of normal for age, and neutrophil count of less than 1500 cells/μL. For patients with interstitial lung disease, this included pulmonary function tests with a forced vital capacity (FVC), total lung capacity (TLC), or diffusion capacity for carbon monoxide less than 80% predicted or a decline of at least 10% in FVC or TLC since the last test.^21^ For patients with immune hepatitis, active disease was defined as the presence of transaminitis and/or positive clinical autoantibody tests (anti-nuclear antigen, anti-smooth muscle actin, anti-liver kidney microsome type 1 and anti-liver cytosol 1).^22^ For patients with thyroiditis, active disease was defined as abnormal levels of thyroid stimulating hormone and free thyroxine. For patients with inflammatory bowel disease, active disease was defined as presence of symptoms or positive findings on endoscopy. For neurologic disorders, ongoing symptoms related to underlying disease as defined by the treating clinician were considered a marker of active disease. Patients who had received intravenous immunoglobulin (IVIG) within 4 weeks of enrollment or rituximab within 12 months of enrollment were considered to be on treatment based on published kinetics of B cell repopulation in patients with autoimmune disorders.^23,24^ Although ITP and autoimmune neutropenia are considered autoantibody-driven cytopenias, the clinically available antiplatelet and anti-neutrophil autoantibody assays lack sufficient sensitivity and specificity to be used as singular diagnostic tests and are not part of the consensus criteria for diagnosing these conditions.^25–27^ Clinical autoantibody testing is not routinely performed for these patients with immune cytopenias at our center, so this data were not available for our cohort.

### 2.2 Immunophenotyping

To minimize T cell death that can occur during the process of cell isolation,^28^ flow cytometric assessment of T cells was done on 100 µL of whole blood collected in sodium heparin anticoagulant tubes. Cell surface staining was performed using the following antibodies: CD4 (BioLegend #317420), PD-1 (BioLegend, #329907), CXCR5 (BioLegend, #356904), CXCR3 (BioLegend, #353716), and CCR6 (BioLegend, #353434). Similar percentages of cTfm subsets were obtained when anti-CD45RA (BioLegend #304129) was added to exclude naïve T cells. Flow cytometry data were acquired on BD LSRFortessa and were analyzed using FlowJo™ v10.8 Software (BD Life Sciences). The upper limit of normal was defined as the 97·5^th^ percentile of control values for T cell phenotyping, in accordance with Clinical and Laboratory Standards Institute (CLSI) guidelines.^29,30^ Increased percentages of CD4^+^CXCR5^+^PD-1^+^ cTfh cells were defined using a threshold of 12% as previously published.^18^

### 2.3 Autoantibody detection

Sengenics KREX technology (i-OME Discovery array in combination with dual color IgG/IgA detection) was used to profile seroreactivity against 1616 self-antigens with preserved native conformation using 100μL of plasma per patient.^31,32^ Controls for this platform included 20 age-matched individuals from the above-described control cohort as well as a commercially available pooled normal plasma (Sigma) from 50 – 70 individuals without clinical evidence of autoimmunity. Plasma was diluted 1:200 for the assay. Autoantibody binding to antigens was measured using an open format microarray laser scanner (Agilent) at 10 µm resolution, with quantification of intensities using GenePix Pro 7 software (Molecular Devices). Net fluorescence intensities were calculated by subtracting background signaling intensities, followed by log2 transformation, Loess normalization^33^ using the normalizeCyclicLoess() function^34^ from R package limma^35^. Batch-correction was performed using the ComBat() function^36^ from R package sva.^37^ Net normalized intensities are reported in supplementary data 1 and 2. To avoid potential confounding from intravenous immunoglobulin (IVIG), we excluded autoantibodies with significantly increased intensities in patients who received IVIG within 4 weeks of study enrollment compared to patients who did not receive IVIG. High titer autoantibodies were those with a titer exceeding the 97.5^th^ percentile in controls based on Clinical and Laboratory Standards Institute guidelines for establishing the upper limit of normal.^38^ Pathway analysis was performed on high titer of autoantibody targets with Enrichr using the KEGG 2021 database. Pathways with a false discovery rate (FDR) <0.05 were considered significant.

### 2.4 Statistical analysis

Unsupervised hierarchical clustering of autoantibody data was performed using Euclidean distance and the Ward D2 method with the ComplexHeatmap R package.^39,40^ For calculating the sensitivity and specificity of each assay discussed, 95% confidence intervals (CIs) were computed using the Brown-Wilson method. To assess the statistical significance of the test’s accuracy, a Fisher’s exact test was performed to compare proportions, with a two-tailed p-value less than 0.05 considered statistically significant. Mann-Whitney test was used for comparisons between two groups. Kruskal-Wallis and Dunn’s multiple comparisons tests were used for comparisons of more than two groups. Statistical tests were performed using Prism v10.0.0 (GraphPad) or R version 4.3.1. Graphs were created with Prism v10.0.0 and illustrations were created with BioRender.com. To identify clinical variables associated with cTfm skewing, a logistic regression was performed using R Statistical Software (version 4.3.1.) with the mctest package.^41^ The outcome variable was dichotomized based on percentages of cTfm above the 97.5^th^ percentile in controls, with participants classified as having skewed or non-skewed percentages. The clinical variables included in the analysis were: diagnosis of a monogenic disorder, the presence or absence of immunomodulatory treatment, and presence or absence of disease activity. Statistical significance was set at *p* < 0.05

## Results

### Patient characteristics

This study enrolled 175 individuals: 101 patients with PIRD and autoimmunity and 74 controls (**Table 1**). The clinical heterogeneity of this patient cohort reflects the phenotypic variability intrinsic to PIRD^1,42^. Immune cytopenias were the most common clinical feature, affecting 80% of the patients. This included 24 patients with ITP, 22 with autoimmune hemolytic anemia (AIHA), and 35 with Evans syndrome (ES). Additional autoimmune conditions in this cohort included hepatitis, intestinal disease, interstitial lung disease, neurologic disease, and thyroiditis (**Table 1**). There were 28 patients (28%) with a monogenic diagnosis (**Supplementary table 1**). At enrollment, 59 patients (58%) had active disease and 74 (73%) were receiving treatment (**Table 1**). A total of 27 patients had a history of rituximab treatment prior to enrollment, of whom 13 had received it within 12 months before enrollment.

### Patients with PIRD have chemokine receptor-skewed cTfm cells

We previously published that percentages of CD4^+^CXCR5^+^PD-1^+^ cTfh cells exceeding 12% of CD4^+^ T cells have 88% sensitivity and 94% specificity for active, multi-systemic autoimmunity and can normalize with immunomodulatory treatment.^18^ In this cohort, patients with PIRD had significantly higher cTfh than controls (median 7.9% in patients, 5.2% in controls, p<0.0001). In contrast to our prior study, the majority of this cohort (72.2%, 73 patients) had a normal percentage of CD4^+^CXCR5^+^PD-1^+^ cTfh cells **(Fig. 1a)**. These patients had disease features known to be associated with normal cTfh cell percentages:^18^ inactive disease (n=35), autoimmunity affecting a single organ without evidence of multi-systemic disease (n=45), and treatment with immunomodulatory therapies (n=42).

**Figure 1.**
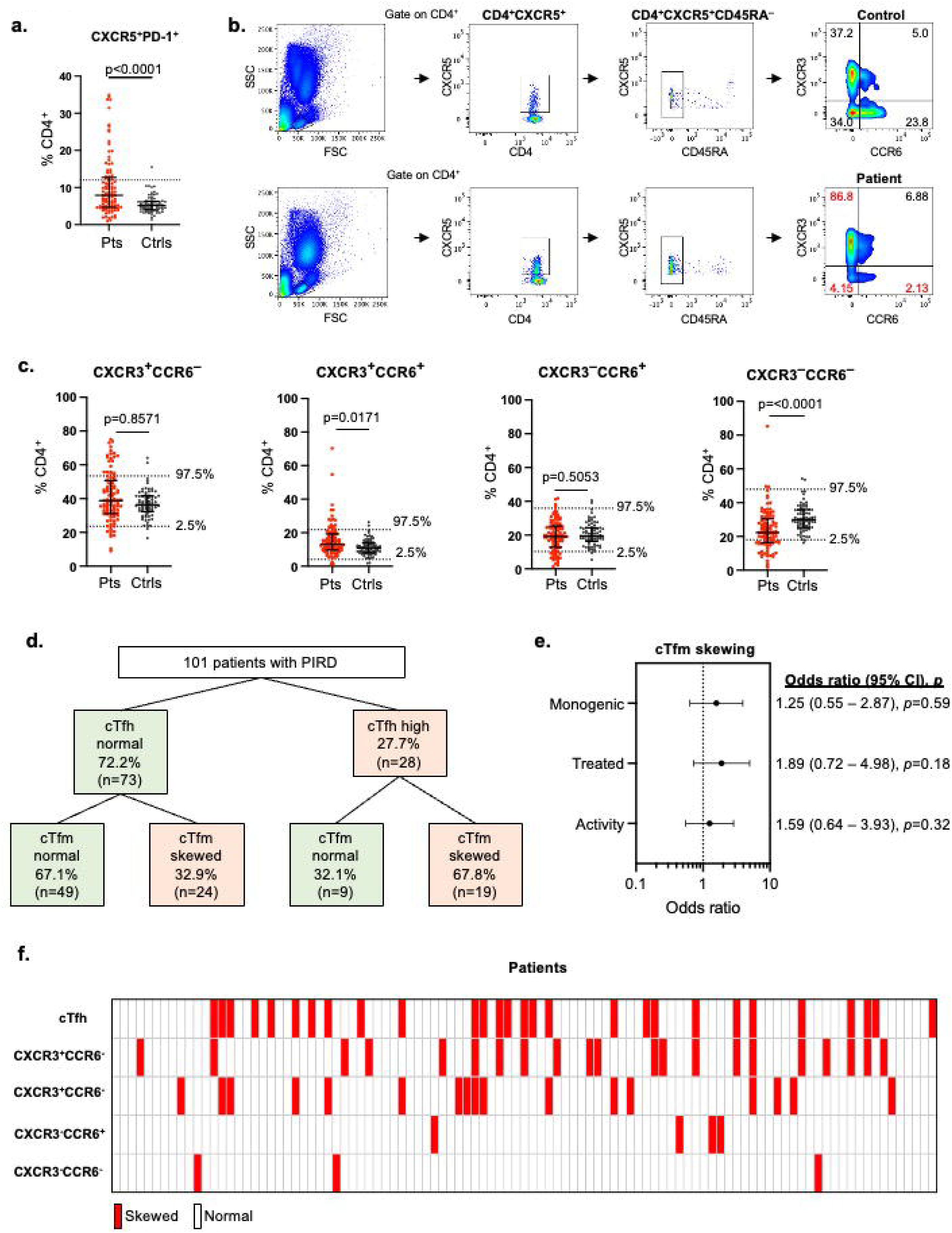
cTfh and cTfm analysis among patients and controls. **a.** Percentages of CD4^+^CXCR5^+^PD-1^+^ cTfh cells in patients (Pts, n=101) and controls (Ctrls, n=74). Dotted line at 12% represents the upper limit of normal, as previously published. *p*-values were calculated by two-tailed Mann-Whitney test. **b.** Gating strategy for identifying chemokine receptor skewing of cTfm cells. **c.** Percentages of CXCR3^+^CCR6^-^, CXCR3^+^CCR6^+^, CXCR3^-^CCR6^+^, CXCR3^-^CCR6^-^ cTfm cells in patients (Pts, n=101) and controls (Ctrls, n=74). *p*-values were calculated by two-tailed Mann-Whitney test adjusted for multiple comparisons. **d.** Flow diagram demonstrating the proportions of patients (n=101) with normal (green) vs. abnormal (red) cTfh % and chemokine receptor skewing of cTfm cells **e.** Multivariate logistic regression of cTfm skewing as a function of presence of monogenic disease, active disease status, and active treatment status. **f.** Distribution of elevated cTfh and chemokine receptor skewing of cTfm cells among individual patients (n=101).

We investigated additional features of antigen-activated CD4^+^ T cells as potential indicators of autoimmunity. We focused on CD4^+^CXCR5^+^ T cells since CXCR5 upregulation occurs after antigen-dependent, cognate interactions essential in autoimmunity.^12^ As CD4^+^ T cells expressing CXCR5^+^ include Tfh cells, cTfh cells, or memory effector cells,^43^ we refer to cells in this early state as circulating T follicular memory (cTfm) cells. We hypothesized that CXCR3 and CCR6 could serve as additional markers of autoimmunity, as these chemokine receptors are upregulated on both cTfh cells and effector memory T cells by inflammatory cytokines and enable recruitment of T cells to inflamed tissues.^14,44^ Prior studies have shown that individuals with a wide range of autoimmune disorders have expansion of cTfh and effector memory T cells expressing different combinations of these chemokine receptors: CXCR3^+^CCR6^−^, CXCR3^+^CCR6^+^, CXCR3^−^CCR6^+^, or CXCR3^−^CCR6^−^.^11,45^ Since the proinflammatory cytokines driving CXCR3 and CCR6 fluctuate dynamically during the disease course of autoimmunity, we investigated chemokine receptor skewing of cTfm cells as an indicator of autoimmunity (**Figure 1b**). We defined chemokine-receptor cTfm skewing as percentages of CXCR3^+^ and/or CCR6^+^ cTfm exceeding the 97.5^th^ percentile of those found in controls (**Figure 1c**), a threshold used in clinical laboratory standards for the upper limit of normal.^38^ Among 101 patients with PIRD 43% of patients had skewed cTfm cells. CXCR3^+^CCR6^−^ and CXCR3^+^CCR6^+^ cTfm cells were the most commonly increased populations in patients, leading to reciprocally reduced percentages of CXCR3^−^CCR6^−^ cTfm cells (**Figure 1c**). Of the 73 patients with normal cTfh, 24 (32.9%) had skewed cTfm cells (**Figure 1d**). cTfm skewing did not differ based on disease activity (*p*=0.59), treatment status (*p*=0.20), or diagnosis of monogenic disorder (*p*=0.32) by multivariate regression (**Figure 1e**). Neither cTfh nor cTfm skewing differed by type of immunomodulatory therapy (**Supplementary Figure 1**). Most patients had non-overlapping patterns of cTfm skewing (**Figure 1f**). Prior studies have identified increased percentages of circulating CD4^+^CXCR5^−^PD1^high^ T peripheral helper (Tph) cells in patients with different types of autoimmune diseases, particularly in common autoimmune disorders, such as rheumatoid arthritis. In our cohort, patients with PIRD had higher percentages of Tph cells compared to controls, with 15 patients having a Tph cell percentage exceeding the 97.5^th^ percentile of controls (**Supplementary Figure 2**). Of these 15 patients, 14 had skewed cTfm cells.

### Patients with PIRD have a high burden of autoantibodies

To complement measures of T cell activation, we investigated autoantibody profiles in a representative sub-cohort of 56 patients with PIRD and 20 controls from our total study cohort. We used an analytic approach that addressed specific challenges hindering the use of autoantibody profiling as a diagnostic tool. First, very few pathogenic autoantibodies have been identified for autoimmune diseases, even for those treated with B cell depleting agents.^7^ Additionally, pediatric patients with PIRD are typically too few in number to power studies characterizing autoantibody repertoires for specific diseases. To circumvent these limitations, we hypothesized that patients with PIRD would have an increased total number and/or diversity of autoantibodies compared to controls, regardless of disease sub-phenotype or autoantibody target. To do this, we used a platform that detects IgG and IgA autoantibodies to 1616 conformationally intact, full-length human proteins related to autoimmunity and cancer.^31,32^ High-titer autoantibodies were classified as those with normalized intensities exceeding three standard deviations of those found in controls. This approach identified a high-titer anti-muscle-specific tyrosine kinase in a patient with myasthenia gravis in the setting of Kabuki syndrome, concordant with clinical testing for autoantibodies in this patient. The targets of high-titer IgG and IgA autoantibodies were predominantly intracellular antigens, likely reflecting antigen release after cell death (**Supplementary Data 3 and 4**).^46^ We defined a high autoantibody burden as a total number of high-titer autoantibodies exceeding the 97.5 percentile of those in controls, which was 15 for IgG and 4 for IgA. Patients had a median of five high-titer IgG autoantibodies (range: 0 – 249 autoantibodies per patient) compared to a median of 1.5 high-titer IgG autoantibodies in controls (range: 0 – 16 autoantibodies per control, **Figure 2a**). There were 530 high-titer IgG autoantibodies found only in the patients with PIRD (**Supplementary Data 3**), whose targets were significantly enriched in pathways pertaining to cell cycling, cell senescence, insulin signaling, mitophagy, and PI3K/AKT/mTOR, a metabolic pathway important for cTfh and effector T cell dysregulation as well as autoimmune cytopenias (**Figure 2b**).^47,48^ Among the eight patients with high IgG autoantibody burden, two had received IVIG within the four weeks preceding enrollment. The remaining 15 patients who had received IVIG within the four weeks prior to enrollment had normal IgG autoantibody burdens. Therefore, patients who received IVIG therapy were not overrepresented among those with a high autoantibody burden. In addition, pathway analysis of high titer-IgG autoantibodies did not differ when IVIG recipients were excluded from the analysis (**Supplementary Data 5**). Among the 32 patients with active disease, six had high IgG autoantibody burden. There were eight patients with active disease who were not on immunomodulatory treatment at the time of enrollment, only one of whom had an elevated IgG autoantibody burden.

**Figure 2.**
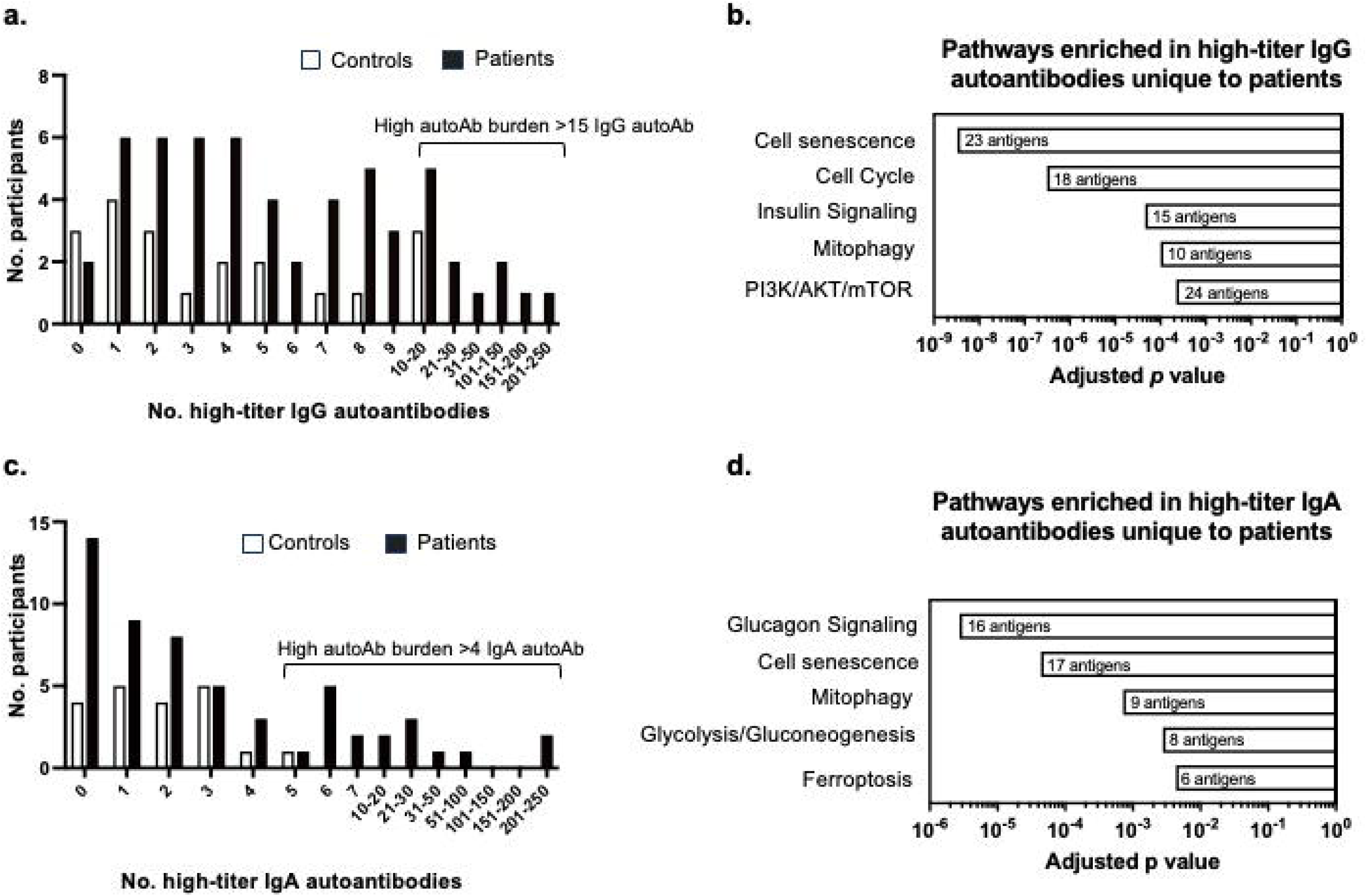
Autoantibody burden and pathway analysis among patients and controls. **a.** Total number of high-titer IgG autoantibodies in patients (black, n=56) and controls (white, n=20). **b.** Pathways enriched in high-titer IgG autoantibodies found only in the patient cohort. **c.** Total number of high-titer IgA autoantibodies in patients (black, n=56) and controls (white, n=20). **d.** Pathways enriched in high-titer IgA autoantibodies found only in the patient cohort.

Patients had a median of two high-titer IgA autoantibodies (range: 0 – 232) and increased autoantibody diversity as compared to a median of one high-titer IgA autoantibody in controls (range: 0 – 5, **Figure 2c**). There were 526 high-titer IgA autoantibodies found only in the patients with autoimmunity (**Supplementary Data 4**), whose targets were significantly enriched in pathways pertaining to cellular senescence, mitophagy, ferroptosis, and glucose metabolism (**Figure 2d**). These pathways have been previously shown to contribute to autoimmunity and overlap with those enriched in targets of high-titer IgG autoantibodies.^49,50^ There were 17 patients with high IgA autoantibody burden, ten of whom had active disease. Among the eight patients with active disease who were not on immunomodulatory treatment at the time of enrollment, two had an elevated IgA autoantibody burden. In this cross-sectional study, the number of high-titer IgG or IgA autoantibodies was not different in patients with active disease or ongoing treatment (**Supplementary Figure 3**). Autoantibody burden was not significantly associated with cTfh elevation (p=0.7715) or cTfm skewing (p=0.5911, **Figure 3a**).

**Figure 3.**
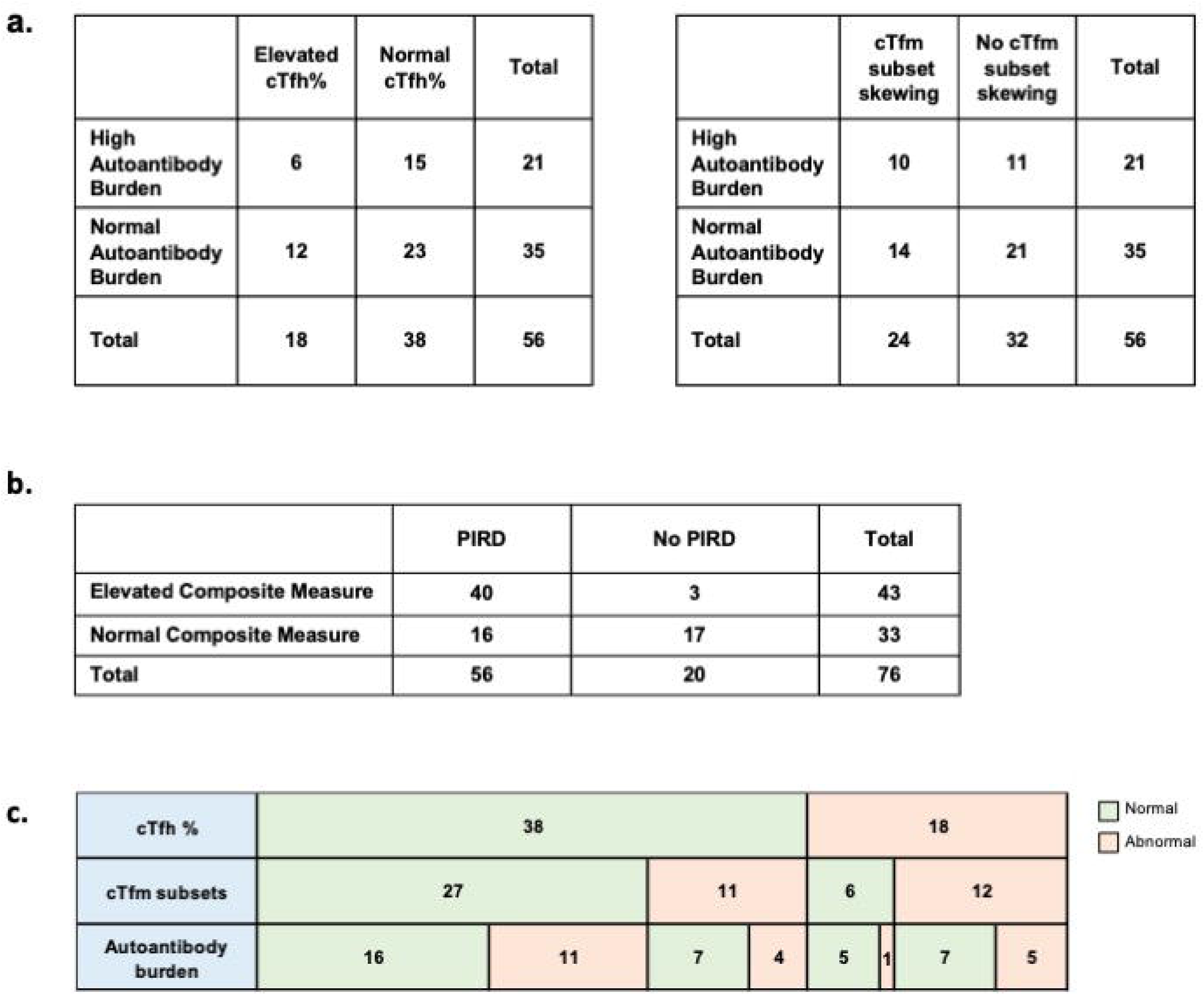
Associations between cTfh%, cTfm skewing, and autoantibody burden. **a**. Contingency tables using Fisher’s exact test to determine that neither elevated cTfh nor cTfm skewing correlates with autoantibody burden. **b.** Contingency table of elevated composite measure, defined as elevated cTfh, cTfm subset skewing, and/or high autoantibody burden in patients with and without PIRD. Sensitivity was defined as the proportion with an elevated composite measure among those who had PIRD (40/56), and specificity was defined as the proportion with a normal composite measure among those who did not have PIRD (17/20)**. c.** Proportion of normal (green) versus abnormal (red) cTfh percentages, cTfm subsets, and autoantibody burdens in cohort of 56 patients with PIRD.

To determine if any specific autoantibodies clustered with clinical phenotypes, we performed unsupervised hierarchical clustering of all autoantibodies in patients and controls. Although we did not find an association between IVIG and the total number of high-titers autoantibodies, IVIG is known to confound analyses of specific autoantibodies in patient populations.^51,52^ Therefore, we eliminated patients who received IVIG within four weeks of study enrollment from the unsupervised hierarchical clustering of autoantibodies (**Supplementary Figure 4**). This analysis did not identify any specific autoantibodies associated with clinical phenotypes (**Supplementary Figure 4**). Unsupervised hierarchical clustering of IgA autoantibodies also did not identify any discernable pattern of autoantibodies associated with clinical features (**Supplementary Figure 5**). Collectively, these findings show that quantification of the autoantibody burden may provide an alternative approach for identifying autoimmunity in patients lacking specific autoantibody targets.

### Combining cTfh, cTfm skewing, and autoantibody profiling for the diagnosis of autoimmunity

Based on the dual contributions of T and B cells in the development of autoimmunity, we tested the sensitivity and specificity of a composite measure integrating increased percentages of cTfh cells and chemokine receptor-skewed cTfm, and a high burden of high-titer IgG and/or IgA autoantibodies for autoimmunity within the cohort of 56 patients and 22 controls for whom all measurements were available. cTfh elevation had sensitivity of 32.1% (95% CI: 0.2410 – 0.4518) and specificity of 100% (95% CI: 0.8389 – 1.0000) for autoimmunity. cTfm skewing had sensitivity of 42.9% (95% CI: 0.3077 – 0.5586) and specificity of 95.0% (95% CI: 0.7639 – 0.9974) for autoimmunity and identified an additional 11 of 38 of patients who had normal percentages of cTfh cells. High IgG or IgA autoantibody burden had sensitivity of 37.5% (95% CI: 0.2601 – 0.5059) and specificity of 90.0% (95% CI: 0.6990 – 0.9822) for autoimmunity and identified an additional 11 of 27 patients who had normal percentages of cTfh and no cTfm skewing. The composite measure of cTfh elevation, cTfm subset skewing, and increased autoantibody burden had sensitivity of 71.4% (95% CI: 0.5852 – 0.8158) and specificity of 85.0% (95% CI: 0.6396 – 0.9476) for autoimmunity in this cohort of patients. (**Figure 3b, c**).

## Discussion

Here, we demonstrate new approaches for characterizing patients with PIRD. Prior studies have identified increased percentages of CD4^+^ T cells expressing CXCR3 and/or CCR6 in different diseases, leading to apparent discrepancies in the subtypes associated with different diseases.^53^ As upregulation of these chemokine receptors is influenced by dynamic changes in inflammatory cytokines inherent in PIRD, we show that chemokine receptor skewing itself is a feature that can be used to detect autoimmunity. The quantification of chemokine receptor-skewed CD4^+^CXCR5^+^ cells uses equipment and techniques readily available in most clinical laboratories with a low reagent cost.^54^ Although the reagent cost of a diverse autoantibody array is more substantial^55^, this assay uses a commercially available protein microarrays and an open-format microarray scanner available in clinical laboratories. This approach complements clinically available measurements of specific antibodies as it can assess serologic autoimmunity even when known pathogenic autoantibodies are unknown or undetectable.

The diversity of clinical phenotypes, disease activity, and therapies in our patient cohort reflects the clinical reality of PIRD. For example, the differential diagnosis for isolated thrombocytopenia includes ITP as well as many other non-autoimmune causes, including familial thrombocytopenia, bone marrow failure syndromes, medication-induced thrombocytopenia, and platelet sequestration in the liver or spleen.^25,27,56^ ITP remains a diagnosis of exclusion, as there are no laboratory tests for diagnosing ITP and current clinical tests for anti-platelet glycoprotein autoantibodies have 53% sensitivity for ITP. Even at centers with a large volume of immune cytopenia cases, up to 14% of children and 12.2% of adults who were diagnosed with ITP were misdiagnosed, while 3.1% of patients diagnosed with other causes of thrombocytopenia were ultimately found to have ITP. ^25,27,56^ Similarly, autoimmune neutropenia is a diagnostic challenge due to the lack of a diagnostic tests specific for autoimmune neutropenia.^26^ Notably, we were able to detect increased percentages of cTfm skewing even in patients with ongoing immunomodulatory treatment. Therefore, a composite measure based on cellular and humoral mechanisms of autoimmunity, rather than disease-specific antigens, may address these diagnostic challenges.

We previously showed that percentages of circulating CD4^+^CXCR5^+^PD1^+^ cTfh cells exceeding 12% of CD4^+^ T cells have high sensitivity and specificity for active, multi-systemic autoimmunity.^18^ Unlike our current study, the prior patient cohort had autoimmunity affecting more than one organ system. Our work complements published studies showing increased cTfh levels in patients with specific autoimmune disorders, such as SLE, rheumatoid arthritis, and Sjogren syndrome.^11,19^ Less is known about the dysregulation of cTfh cells in patients with autoimmune cytopenias, which affected 84% of the patients in our study. In a cohort of 24 patients with Evans syndrome due to the presence of at least two types of autoimmune cytopenias and 22 patients with chronic ITP, Kumar et al. found that patients with Evans Syndrome have increased percentages of cTfh cells and CXCR3^+^CXCR5^+^PD-1^+^ T cells compared to those with chronic ITP.^57^ While this prior study focused on the CD4^+^CXCR5^+^PD-1^+^ T cells, we quantified total CD4^+^CXCR5^+^ T cells regardless of PD-1 expression. We chose this approach because CD4^+^CXCR5^+^ T cells lacking PD-1 can also contribute to autoimmunity by eventually differentiating into effector memory T cells expressing either CXCR3 or CCR6.^58^ Measurement of CD4^+^CXCR5^+^ cTfm cells thus encompasses different types of antigen-experienced T cell populations with distinct contributions to autoimmunity. The increased expression of CXCR3^+^ on cTfm cells in our patient cohort (**Figure 1C**) may be arising at least in part from the known roles of IFN-γ in upregulating CXCR3 and in driving autoimmunity.^59–61^ We also show that all but one of the patients with increased Tph cells also had chemokine-skewed cTfm cells. Although these two populations are differentiated by the absence (Tph) or presence (cTfm) of CXCR5^+^, the expansion of Tph cells, like upregulation of CXCR3 and CCR6,^14,44^ is driven by inflammatory cytokines.^62^ We did not integrate Tph cells with other measures of autoimmunity because Rao *et al.* have noted that these cells are infrequent in blood and that studies of Tph cells enriched in inflamed tissue sites, rather than those circulating in the periphery, are the most informative.^63^

We found that percentages of chemokine receptor-skewed cTfm cells did not correlate with whether patients were receiving ongoing immunomodulatory treatment. The most common treatments used in this cohort were glucocorticoids and rituximab. This is clinically relevant since empiric immunosuppressive therapy is sometimes required for management of acute and/or severe autoimmunity before the diagnostic evaluation is complete. Thus, measurement of chemokine receptor-skewed cTfm cells may serve as an indicator of autoimmunity even if other inflammatory markers are normalized by immunomodulatory therapies. Effector T cells expressing CXCR3 and CCR6 have been shown to resist glucocorticoid-induced cell death due to the expression of the multi-drug resistance type 1 membrane transporter, which enable excretion of glucocorticoids, and low expression of the glucocorticoid receptor, further reducing signaling downstream of glucocorticoids.^64,65^ We hypothesize that these mechanisms may underlie the resistance of cTfm cells to treatment, but additional studies are needed to determine this.

We complemented these studies of cTfm subsets with a broad autoantibody profiling to assess the overall burden of high-titer autoantibodies since disease-associated autoantibodies are unknown for many autoimmune disorders. Even for disorders with known autoantibodies, the relationship between autoantibodies and disease activity is unpredictable, since autoantibodies can develop before clinical manifestations of autoimmune diseases.^2^ Our pediatric patient cohort with PIRD harbored a diverse set of high-titer IgG and IgA autoantibodies not found in controls, with more patients having an increased number of high-titer IgA than IgG autoantibodies. The high-titer IgA autoantibodies unique to the patients targeted self-antigens enriched in pathways similar to those targeted by the IgG autoantibodies, suggesting that screening for IgA autoantibodies may comprise another readout for some patients with PIRD. Measuring IgA autoantibodies also circumvents potential confounding from IVIG, although we found no correlation between IVIG treatment and an increased total autoantibody burden. Pathway analysis for enriched high titer-IgG antibodies was also similar regardless of IVIG status. We did not find an association between cTfh cells and the numbers of high-titer autoantibodies, consistent with studies showing that additional factors, including subtypes of B cells and the cytokine milieu, are needed for the generation of autoantibodies. ^2^

This study has several limitations. This study did not include patients with clinical or laboratory evidence of a concomitant infection since infections affect many features of autoimmune diseases, including percentages of cTfh cells and effector memory T cells, inflammatory markers, and cytokine levels.^66–68^ Most of the patients in this study had active disease at the time of enrollment, reflecting the chronic and persistent nature of these disorders. Future longitudinal studies are needed to delineate how cTfm and autoantibody profiles change through a patient’s disease course. Thus, the clinical context is essential for interpreting these studies. Our study was not powered to identify specific autoantibodies associated with disease phenotypes. Rather, we identified features that can be used to characterize PIRD, which are often diseases of exclusion, without known pathogenic autoantibodies or cellular markers of autoimmunity. Further studies are needed to determine if the measures investigated in this study are generalizable to cohorts from other countries, genetic backgrounds, older age groups, and more common autoimmune diseases.

In conclusion, this study applied two consequences of sustained T-B cell crosstalk to identify new approaches for characterizing diverse types of autoimmunity. We show that chemokine skewing of CD4^+^CXCR5^+^ cTfm cell subsets and quantification of the total burden of autoantibodies can help identify autoimmunity even when percentages of cTfh cells are normal. These approaches thus present new avenues for improved characterization of rare and diverse disorders of autoimmunity.

## Contributor Statement

EMH, SC, BL, and JC conceptualized the study. AC, MN, AB, LM, ME, and BL obtained consent, processed patient samples, performed experiments, and contributed to data collection and analysis. LS, MG-A, TKO, JMP, EMH, SC, and JC verified and analyzed the data and generated figures. EMH, SC, DPH, CDP, RFG, BL, and JC contributed to patient phenotyping. EMH, SC, and JC wrote the initial version of the manuscript. AA-M, BW, EO, SS, DPH, CSKY, RWN, PL, OH, RCH, MD-L, LAH, AAN,CDP, and RFG provided feedback and revised the manuscript. All authors have read and approved the final version.

## Data Sharing Statement

Deidentified data will be provided to qualified investigators upon reasonable request to the corresponding author.

## Data Availability

Research data will be made available upon request to the authors

## Abbreviations

AIHA: Autoimmune hemolytic anemia
cTfh: Circulating T follicular helper cell
cTfm: Circulating T follicular memory cell
ES: Evans syndrome
ITP: Immune thrombocytopenia
IVIG: Intravenous immunoglobulin
PIRD: Primary immune regulatory disorders
SLE: Systemic lupus erythematosus

## Acknowledgments

We are grateful to our patients and their families for their participation in this study.

## Figure legends

**Supplementary Figure 1. Percentages of cTfh cells and cTfm cells in patients receiving different types of treatments.** For each treatment, “+” and “–” indicates that the patient was either receiving or not receiving each treatment, respectively, as described in the methods. **a.** IVIG (+ n=29, – n=72) **b.** Corticosteroids (+ n=23, – n=78) **c.** Rituximab (+ n=13, – n=88) **d.** Mycophenolate (+ n=15, – n=86) **e.** Sirolimus (+ n=9, – n=92). Horizontal dotted line at 12% on cTfh plots represents the upper limit of normal, as previously published. Horizontal dotted lines on cTfm plots represent 97.5 and 2.5 percentile of controls. *p*-values were calculated by two-tailed Mann-Whitney test adjusted for multiple comparisons.

**Supplementary Figure 2. Percentages of CD4^+^CXCR5^−^PD1^high^ Tph cells in patients (Pts) and controls (Ctrls) \*\***p<0.01 by two-tailed Mann-Whitney test.

**Supplementary Figure 3. Association of high-titer autoantibody number with disease activity and treatment status**. **a.** Number of high-titer IgG and IgA autoantibodies in patients with active (+) and inactive (-) disease. **b.** Number of high-titer IgG and IgA autoantibodies in patients on treatment (+) and not on treatment (-) at time of enrollment. Horizontal lines indicate median, error bars indicate the interquartile range. ns, *p*>0.05 by two-tailed Mann-Whitney test. Y axis is log_2_ scaled. UD = undetectable

**Supplementary Figure 4. Heatmap with unsupervised clustering based on the normalized net intensities of IgG autoantibodies in patients and controls.** Patients who had received IVIG within the preceding four weeks were excluded. Red: present, grey: inactive, white: not present (control).

**Supplementary Figure 5. Heatmap with unsupervised clustering based on the normalized net intensities of IgA autoantibodies in patients and controls.** Red: present, grey: inactive, white: not present (control).

**Supplementary Table 1:** Genetic diagnoses of patients with monogenic disorders

**Supplementary Data 1.** IgG autoantibody net normalized intensities in patients and controls

**Supplementary Data 2.** IgA autoantibody net normalized intensities in patients and controls

**Supplementary Data 3.** High titer IgG autoantibodies with subcellular locations

**Supplementary Data 4:** High titer IgA autoantibodies with subcellular locations

**Supplementary Data 5:** Comparison of the top five enriched pathways for IgG targets in cohort including IVIg recipients (n=56) as compared to cohort excluding IVIg recipients (n=39).

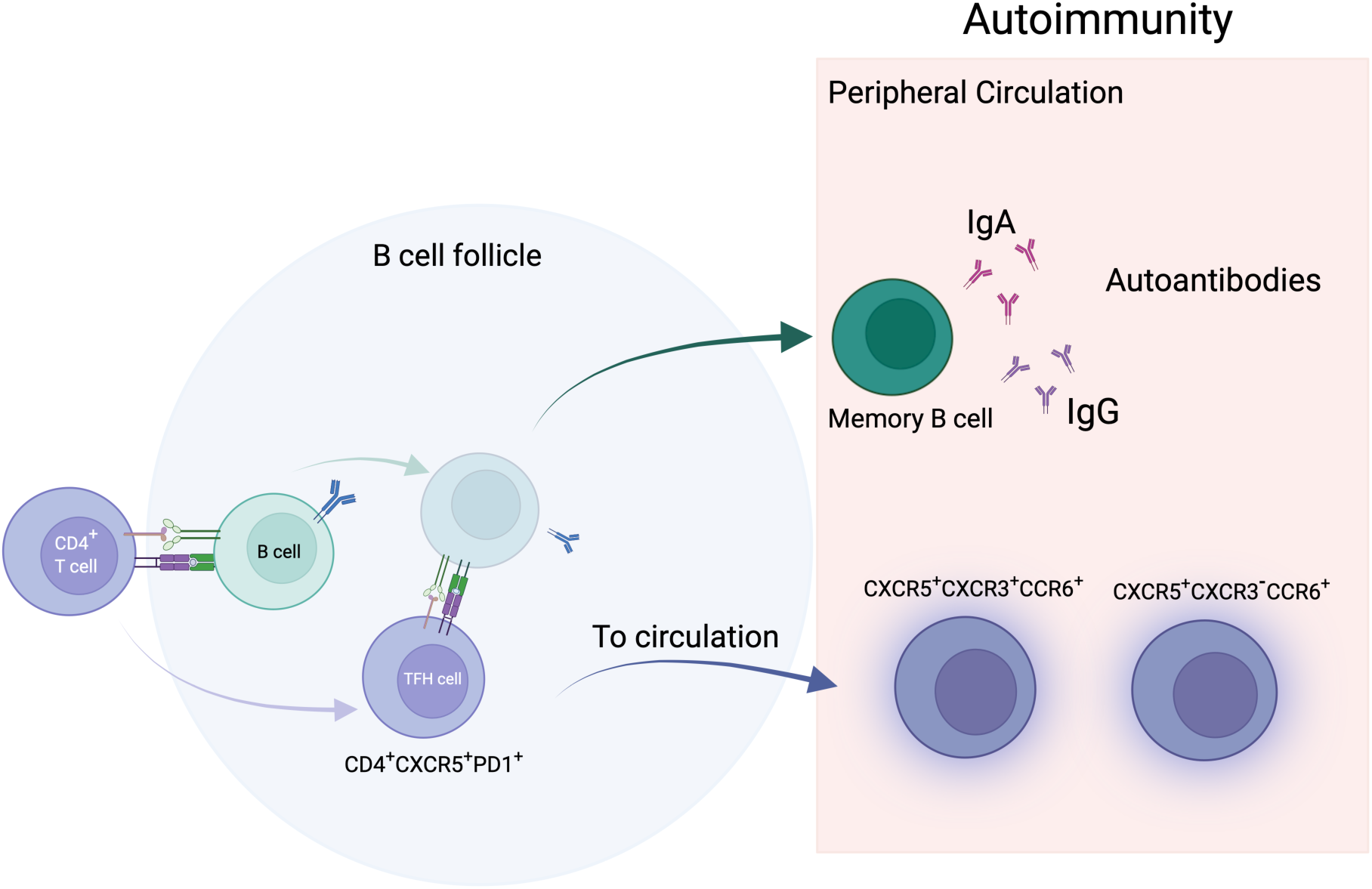

