## Supplementary figures and images for "T cell and autoantibody profiling for primary immune regulatory disorders"

### Supplementary Figure 1

#### Slide 1
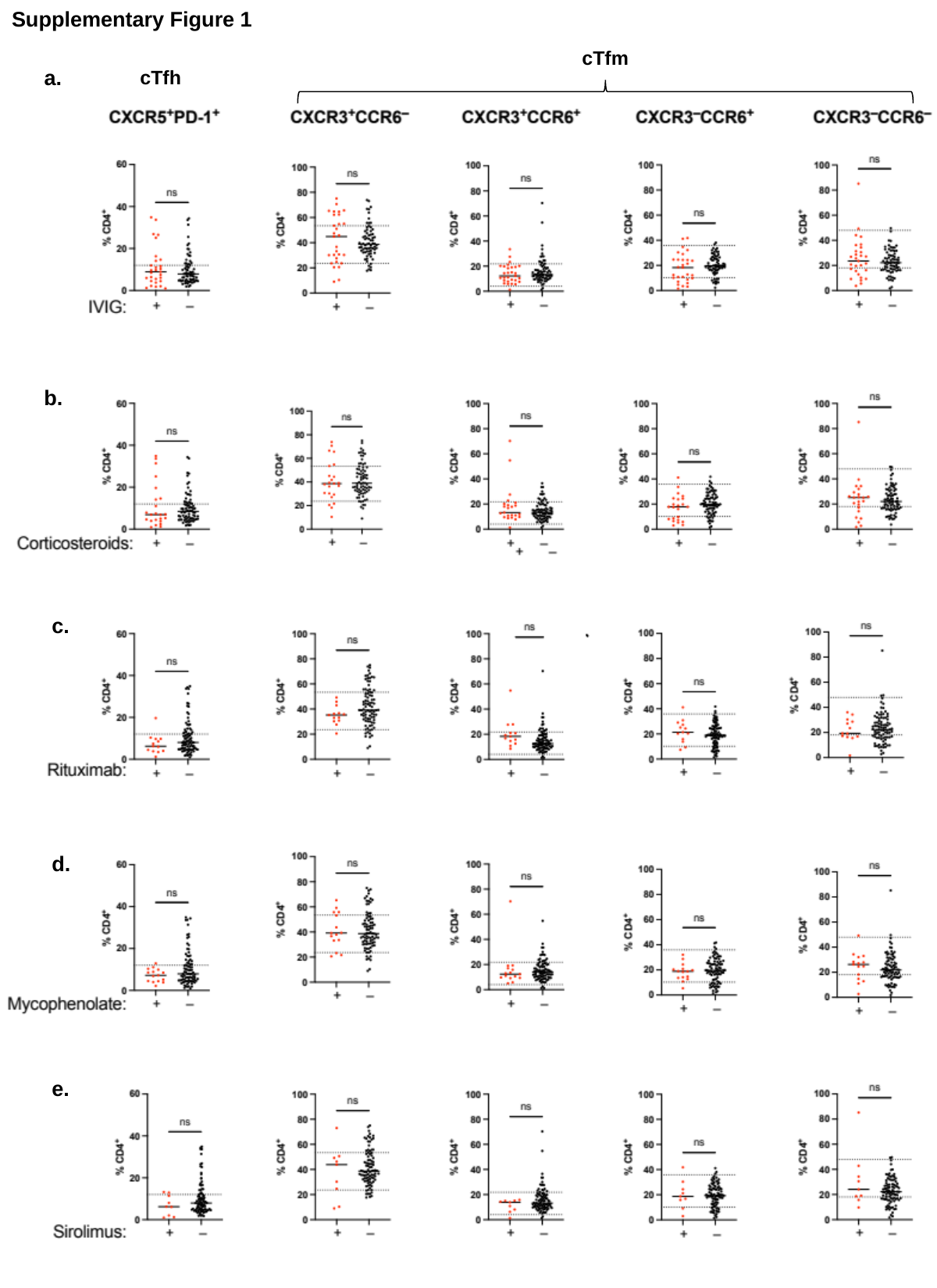

Supplementary Figure 1
cTfm
a.
cTfh
b.
c.
d.
e.

### Supplementary Figure 2

#### Slide 1
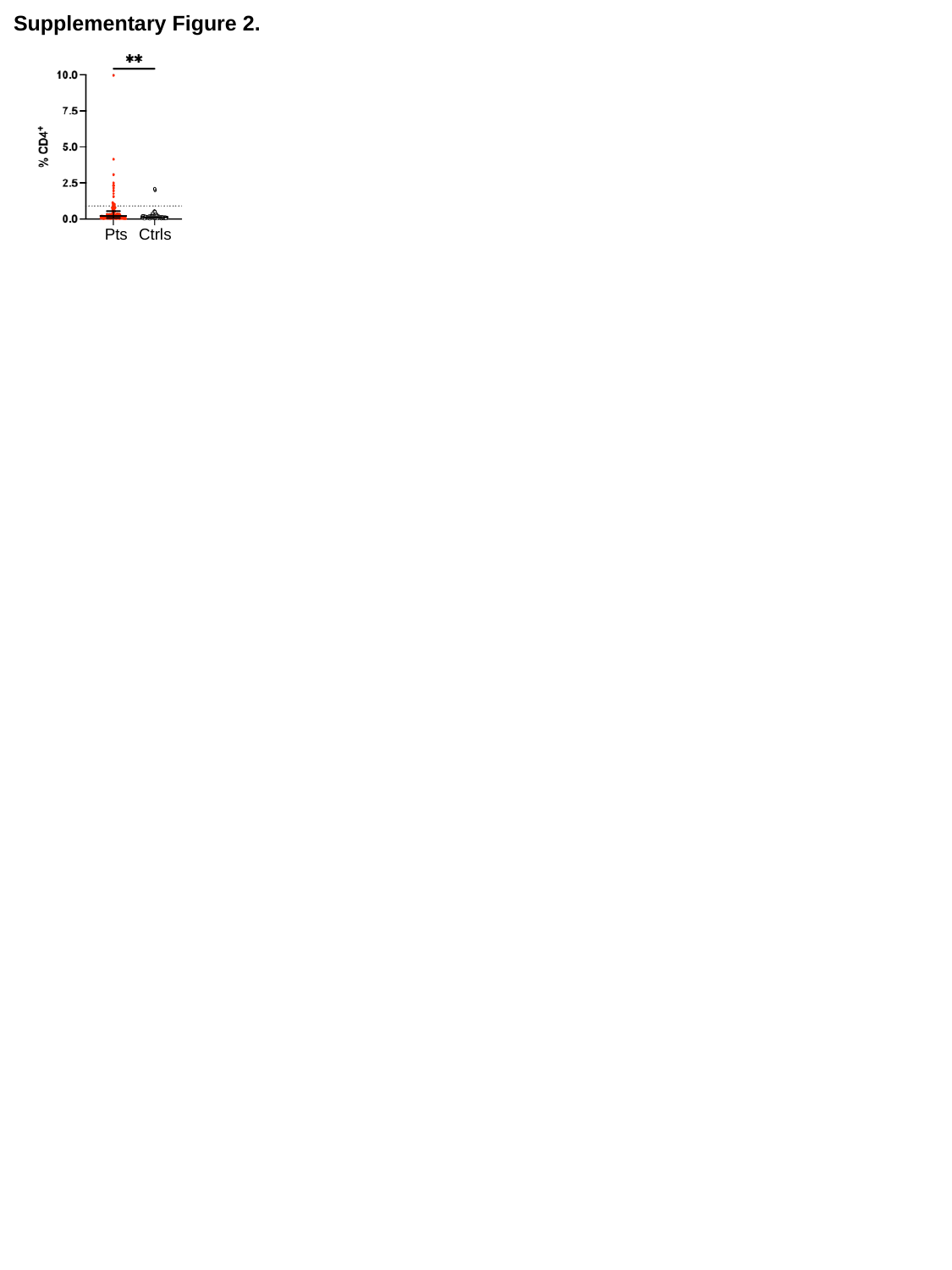

Supplementary Figure 2.
Pts
Ctrls

### Supplementary Figure 3

#### Slide 1
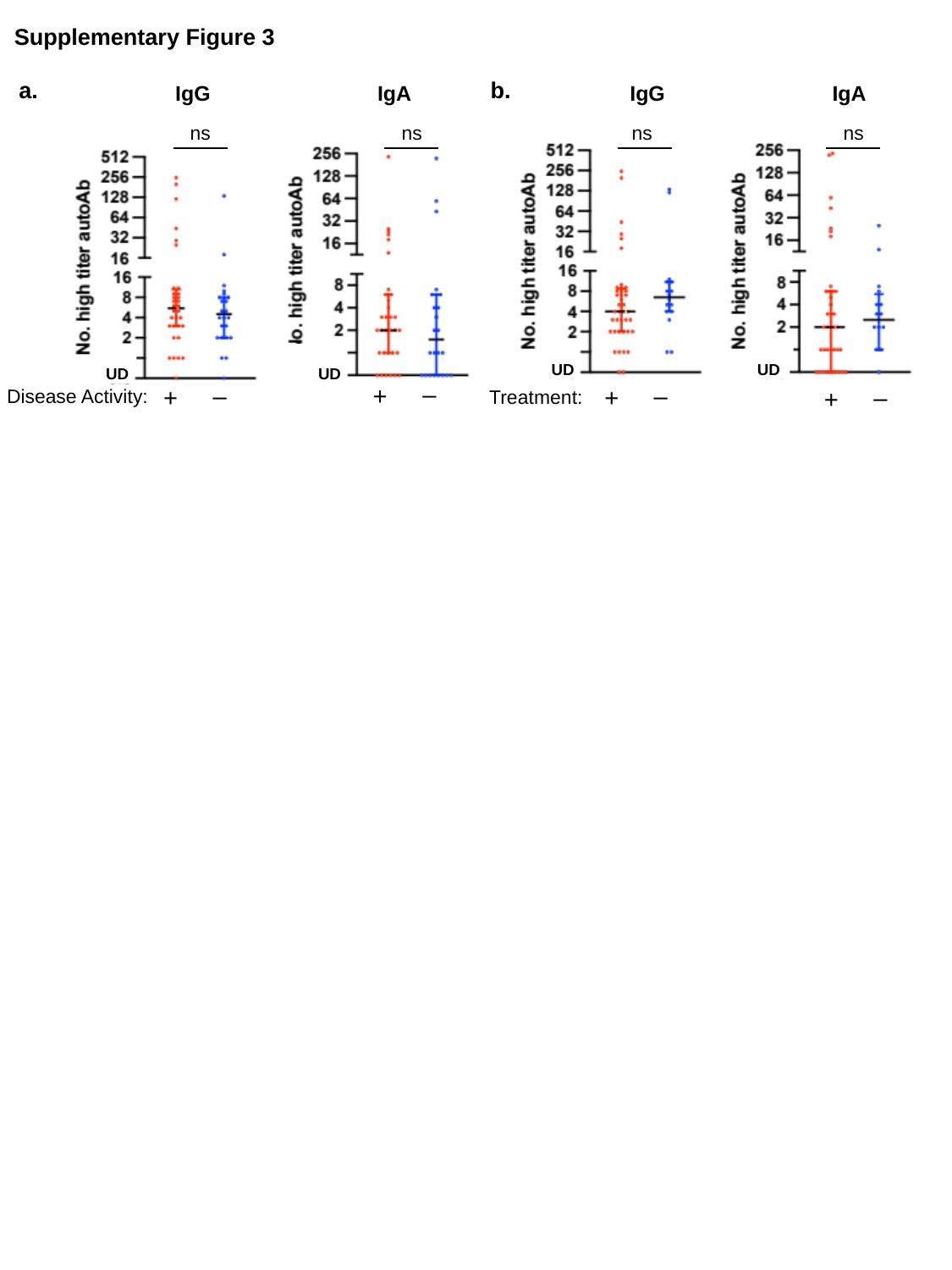

Supplementary Figure 3
a.
b.
IgG
IgA
IgG
IgA
ns
ns
ns
ns
UD
UD
UD
UD
–
+
–
+
–
+
–
+
Disease Activity:
Treatment:
