## Supplementary Figure 4 for "T cell and autoantibody profiling for primary immune regulatory disorders"

### Slide 1
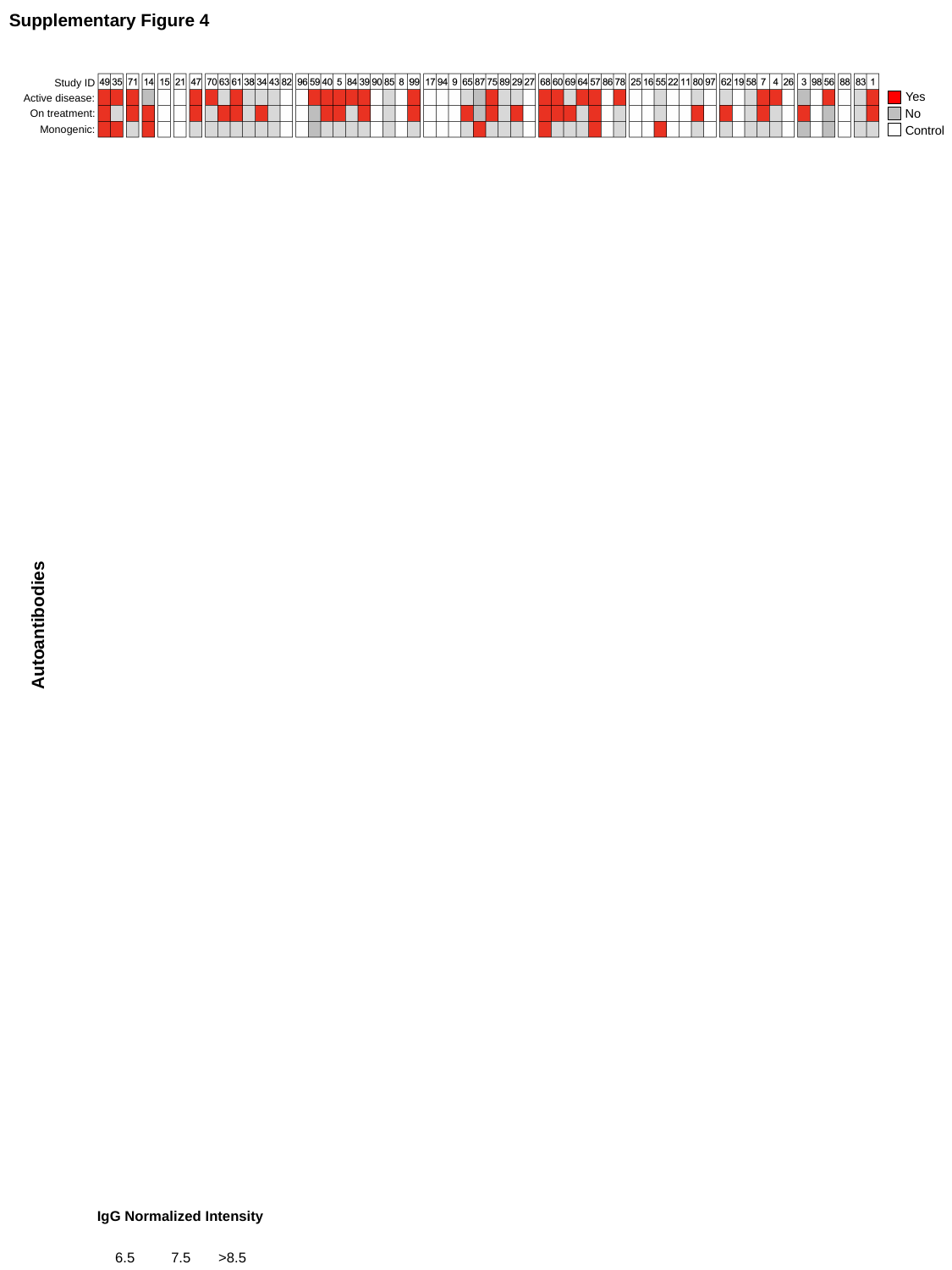

Supplementary Figure 4
Study ID
Yes
Active disease:
No
On treatment:
Control
Monogenic:
Autoantibodies
IgG Normalized Intensity
6.5
7.5
>8.5
