## Supplementary Figure 5 for "T cell and autoantibody profiling for primary immune regulatory disorders"

### Slide 1
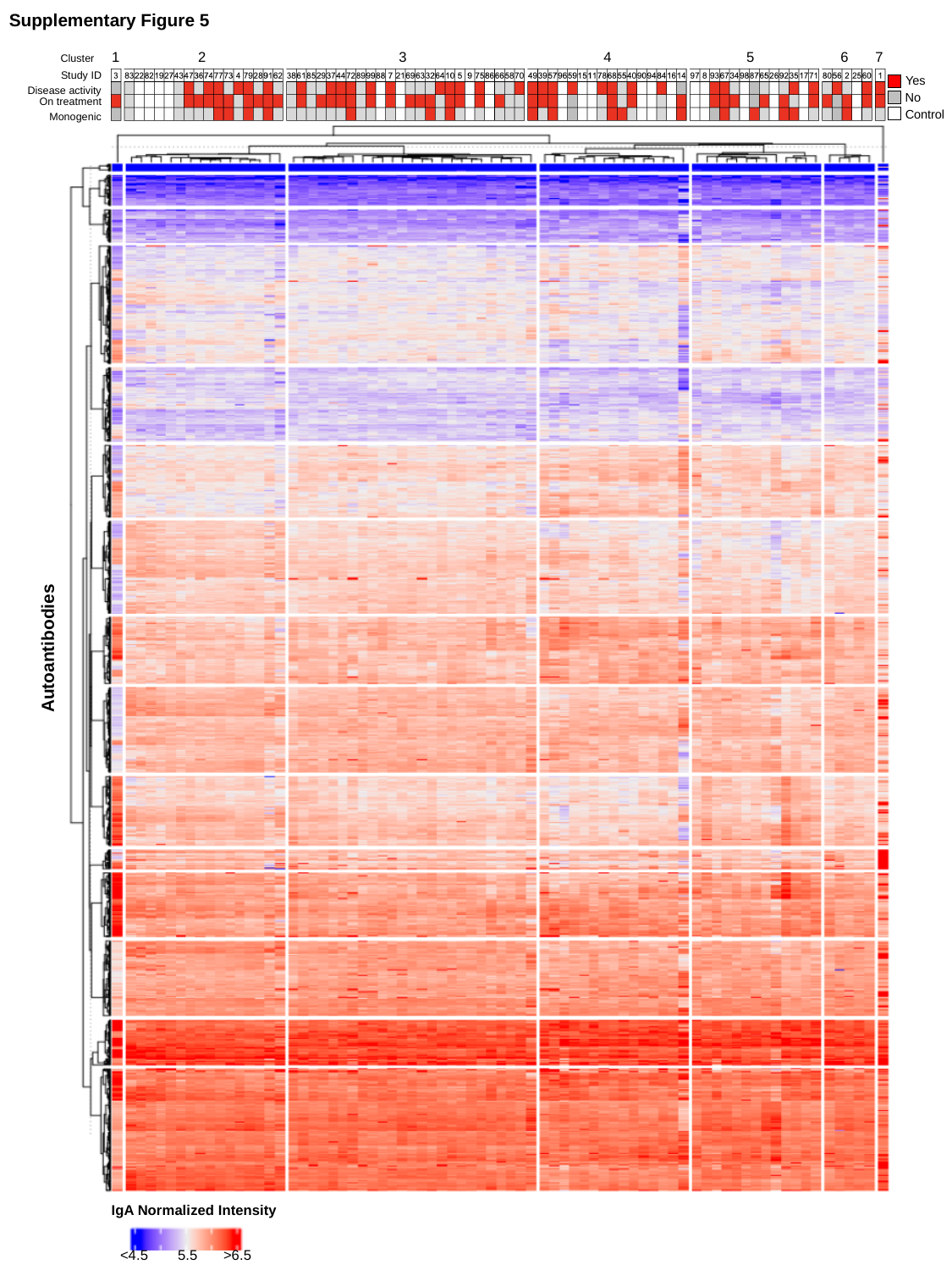

Supplementary Figure 5
1
2
3
4
5
6
7
Cluster
Study ID
Yes
Disease activity
No
On treatment
Control
Monogenic
Autoantibodies
IgA Normalized Intensity
<4.5
5.5
>6.5
