## Supplementary Table I for "T cell and autoantibody profiling for primary immune regulatory disorders"

**Supplementary Table I**. List of genetic diagnoses in patients with monogenic disorders. All variants are heterozygous unless otherwise indicated.

| **Monogenic Disorder** | **Gene** | **Number of patients** | **Causative variants** |
| --- | --- | --- | --- |
| ALPS | *FAS* | 1 | c.617del, p.Asn206Thrfs*10 |
| APS-1 | *AIRE* | 1 | c.1370G>A, p.Cys457Tyr (biallelic) |
| APDS | *PI3KCD* | 1 | c.3061G>A, p.Glu1021Lys |
| BACH2 deficiency | *BACH2* | 1 | c.2230A>G p.Ile744Val (biallelic) |
| COPA syndrome | *COPA* | 1 | c.698G>A, p.Arg233His |
| *CTLA4* haploinsufficiency | *CTLA4* | 4 | c.347T>A, p.Ile116Asn |
|  |  |  | Chromosome 2q33.1q33.3 deletion |
|  |  |  | c.654T>A, p.Tyr218Ter |
|  |  |  | c.654T>A, p.Tyr218Ter |
| Kabuki syndrome | *KMT2D* | 6 | c.12415_12416del, p.Val4139Phefs*28 |
|  |  |  | c.44221G>T, p.Cys1474Phe |
|  |  |  | c.3082delC, p.Leu1028Phefs*X28 |
|  |  |  | c.16267G>T, p.Val5423Phe |
|  |  |  | c.6265_6266delinsCAT |
|  |  |  | c.15731_15732delAA |
| Moesin deficiency | *MSN* | 1 | c.1678C>T, p.Arg560Cys |
| *NFKB1* haploinsufficiency | *NFKB1* | 1 | c.259-1G>C |
|  |  |  | c.884G>A; p.Trp295* |
| *NFKB2 Haploinsufficiency* | *NFKB2* | 1 | c.1354G>A, p.Gly452Ser |
| PLAID | *PLCG2* | 2 | c.2866C>T, p.Arg956Cys |
|  |  |  | c.3420T>A; p.Asp1140Glu |
| RAC2 Gain of Function | *RAC2* | 1 | c.275A >C, p.Asn92Thr |
| RAG1 deficiency | *RAG1* | 1 | c.1210C>T, p.Arg404Trp and c.983G>A, p.Cys328Tyr |
| *SOCS1* haploinsufficiency | *SOCS1* | 1 | c.24delA, p.Ala9Profs∗7 |
| *STAT1* gain-of-function | *STAT1* | 2 | c.604A>G, p.Met202Val |
|  |  |  | c.821G>A, p.Arg274Gly |
| *STAT3* gain-of-function | *STAT3* | 2 | c.1199A>T, p.Asn400Ile |
|  |  |  | c.2144C>T, p.Pro715Leu |

ALPS: Autoimmune lymphoproliferative syndrome;APS-1, Autoimmune polyglandular syndrome type 1; APDS: Activated PI3K delta syndrome; PLAID, PLCG2-associated antibody deficiency and immune dysregulation.
